# Characterization of Congenital Hyperinsulinism in Argentina: Clinical Features, Genetic Findings, and Treatment Outcomes

**DOI:** 10.1101/2025.03.14.25323434

**Authors:** Gabriela Pacheco, Maria G. Bastida, Juan Cáceres, Guillermo Alonso, Mariana Aziz, Martha Suarez, Adriana Flores, Victoria Femenia, María V. Forclaz, Jayne A.L. Houghton, Jasmin J. Bennett, Sabrina Martin, Sarah E. Flanagan, Ana Tangari-Saredo

**Affiliations:** Pediatric Nutrology, Hospital Público Materno Infantil de Salta, Argentina; Hospital Provincial Neuquén, Dr E Castro Rendón, Neuquén, Argentina; Endocrinology, Hospital Plottier, Neuquén, Argentina; Pediatric Endocrinology Section, Hospital Italiano de Buenos Aires, Buenos Aires, Argentina; Department of Endocrinology, Hospital de Pediatría J.P. Garrahan, Buenos Aires, Argentina; Department of Medicine, Endocrinology Unit, Hospital T. Alvarez, Buenos Aires, Argentina; Nutrition and Diabetes Service, Fundación Hospitalaria, Buenos Aires, Argentina; Endocrinology, Hospital H. Notti, Mendoza, Argentina; Pediatric Endocrine Service, Hospital Posadas, Buenos Aires, Argentina; Department of Clinical and Biomedical Science, University of Exeter, Exeter, UK; The Genomics Laboratory, Royal Devon University Healthcare NHS Foundation Trust, Exeter, UK; Division of Endocrinology, Sanatorio Güemes, Buenos Aires, Argentina

## Abstract

**Introduction:** Congenital hyperinsulinism (CHI) is a heterogeneous disorder of insulin dysregulation, leading to hypoglycemia. This study describes the clinical characteristics, genetics, and management of CHI in Argentina.

**Methods:** We retrospectively reviewed 70 probands diagnosed with CHI at multiple centres across Argentina. Clinical, biochemical, imaging, and treatment data were analyzed. Genetic testing was performed in 49 probands using Sanger and targeted next-generation sequencing of CHI-related genes.

**Results:** Transient CHI was identified in 23/70 (33%) probands, with a median duration of 2 months. Risk factors for perinatal stress-induced hyperinsulinism (PSHI) were present in 85% of transient cases. Persistent CHI was diagnosed in 44/70 (63%) individuals, of whom 31 responded to diazoxide. Late-onset CHI (diagnosed >3 years) was identified in 3 children.

A pathogenic variant was detected in 19/49 (39%) probands, all had persistent CHI. *ABCC8* variants were most common accounting for 68% (13/19) of diagnoses. Imaging in 17 cases revealed focal disease in 8, diffuse disease in 8, and atypical disease in 1 individual. Seven individuals with focal disease underwent lesionectomy, which was curative in 5 (71%). Three children with diffuse disease required near-total pancreatectomy, with one developing postoperative diabetes.

**Conclusions:** This study provides the largest CHI cohort reported from South America and highlights the clinical and genetic heterogeneity of the condition. Transient CHI was often associated with PSHI risk factors, while persistent CHI was predominantly linked to K-ATP channel variants. The findings underscore the importance of genetics and imaging for CHI management and emphasize the need for increased access to molecular diagnostics.

## Introduction

Congenital hyperinsulinism (CHI) causes severe, persistent hypoglycemia. It is diagnosed when there is increased insulin action and/or inadequate suppression of plasma insulin during spontaneous or fasting-induced hypoglycaemia.^1^ In some cases, CHI may be secondary to perinatal factors such as intrauterine growth restriction (IUGR), maternal diabetes mellitus, or perinatal asphyxia.^2^ The hypoglycaemia associated with perinatal stress-induced hyperinsulinism (PSHI) typically resolves within the first weeks of life but can persist for several months in severe cases.^3^

CHI that persists beyond six months affects around 1 in 28,000 live births and is often genetic.^4,5^ More than 30 genetic forms of CHI have been reported, causing either isolated or syndromic disease.^6,7^ Loss-of-function variants in the *ABCC8* and *KCNJ11* genes, which encode the pancreatic ATP-sensitive potassium (K-ATP) channel, are the most common cause of isolated CHI, accounting for ∼50% of cases.^8-10^ Other common genetic causes of persistent CHI include *GLUD1* variants,^11^ and variants affecting the hexokinase genes *GCK* and *HK1*.^12,13^

The genetic cause of CHI determines whether pancreatic histology is diffuse, focal, or atypical.^1^ A focal lesion arises when a paternally inherited *ABCC8* or *KCNJ11* pathogenic variant is unmasked by uniparental isodisomy in pancreatic tissue.^14-16^ In focal CHI, imaging is crucial for localising the lesion before lesionectomy, which is curative in up to 97% of cases.^17,18^ Diffuse disease results from biallelic or dominant variants in the CHI genes.

Those with diffuse CHI who are unresponsive to maximal medical therapy, including diazoxide or somatostatin analogues, may require near-total pancreatectomy which carries a high risk of permanent diabetes and exocrine insufficiency.^17,19,20^ Atypical histological disease has been reported in a few individuals with somatic mosaic variants.^21-23^

There is limited data on the genetics and clinical management of CHI in South America. This study aims to describe our experience in managing children with this heterogeneous condition in Argentina.

## Materials and Methods

### Subjects

Clinical and genetic data were retrospectively collected from clinicians managing children diagnosed with CHI at centres across Argentina. Analysis of clinical data was conducted jointly between the consulting centres. CHI was diagnosed based on detectable plasma insulin during hypoglycaemia (<60 mg/dl). When available, additional biochemical markers, including beta-hydroxybutyrate (<1.8 mmol/L) and free fatty acids (<1.7 mmol/L), along with an inappropriate increase in blood glucose (> 30 mg/dL) following parenteral glucagon administration, were used to support the clinical diagnosis of CHI. Individuals with evidence of syndromic disease at the time of CHI diagnosis were excluded.

The study complied with the Declaration of Helsinki, with informed written consent obtained from the parents of all patients. The study was approved by the Wales Research Ethics Committee 5 (22/WA/0268), with participants recruited to the Genetic Beta Cell Research Bank (IRAS: 316050).

Transient CHI was defined as full remission by six months of age, confirmed by cessation of all treatment. CHI persisting beyond six months was classified as persistent and further subdivided based on diazoxide response. Diazoxide-responsive individuals maintained normal plasma glucose levels for at least 4.5 to 6 hours during a fasting test, while those on 15 mg/kg/day of diazoxide who failed to sustain glucose above 70 mg/dL for 6-hours were classified as diazoxide-unresponsive. Late-onset was defined as CHI diagnosed after 3 years.

### Molecular Genetics

Genetic testing was performed on 49 probands. Of these, 43 were tested at the Exeter Genomics Laboratory and six received testing through a commercial laboratory. Of the 49 children who underwent testing, 7 had transient CHI, 39 had persistent CHI and 3 had late-onset CHI.

In Exeter, Sanger sequencing was the first-line test when clinical features suggested a specific genetic subtype (e.g. *GLUD1* in individuals with hyperammonaemia).^11^ For the remaining individuals, and those without variants identified by Sanger sequencing, targeted next-generation sequencing (tNGS) of 15 CHI genes (*KCNJ11, ABCC8, GLUD1, GCK, HADH, HK1, HNF1A/4A, INSR, KDM6A, KMT2D, SLC16A1, CACNA1D, PMM2, TRMT10A*) was performed as previously described.^24^ This included on- and off-target copy number variant analysis.^25^ Parents and clinically affected family members were tested by Sanger sequencing and variants were classified according to established guidelines.^26,27^

### Imaging

^18^F-DOPA PET-CT imaging was performed on a Philips Gemini True Flight Technology 64-detector-row machine. The images were reconstructed in axial, sagittal and coronal projections and interpreted both with and without attenuation corrected. Early abdominal images were acquired at 30 minutes, while late whole-body images were acquired 60 minutes post-injection.

### Statistical analysis

ANOVA was used to compare quantitative variables, and the Kruskal-Wallis test was applied to normally distributed data. *P*-values <0.05 were considered significant.

## Results

A total of 70 probands were identified. Consanguinity was not reported in any of the families. In 23 of 70 (33%) children, CHI remitted before six months, with an average disease duration of 2 months [IQR: 0.5-6 months]. Nineteen of the 23 children (83%) with transient CHI had one or more risk factor(s) for PSHI, including IUGR (n=8), gestational hypertension and preclampsia (n=5), gestational diabetes (n=4), prematurity (gestational age < 37 weeks) (n=6), fetal bradycardia (n=1), or erythroblastosis (n=1). Sixteen children with transient CHI were treated with diazoxide (mean dose 8.4 mg/kg/day) without adverse effects, while 7 were managed with IV glucose and feeding strategies.

CHI was diagnosed in early infancy and persisted beyond six months in 44 of 70 (63%) children. Of these, 31 (70%) responded well to diazoxide. Treatment was discontinued in eight children before the age of 8 years. In the remaining 23, diazoxide dose was reduced from a mean of 10.8 to 7.2 mg/kg/day after a median of 34 months. Adverse effects of diazoxide were observed in four children (receiving 10-12 mg/kg/day), including fluid overload (two without prophylactic diuretics) and, in two cases, neutropenia or thrombocytopenia. In one child, fluid overload resolved after a reduction in the diazoxide dose and the introduction of octreotide therapy. In the remaining three, adjusting the diuretic dose resolved the fluid excess. When patients did not respond to diazoxide, treatment with octreotide was initiated (median dose: 21 ug/kg/day, range: 9-48 ug/kg/day) and/or frequent feeding. One of these children developed gallstones.

CHI was diagnosed in three individuals after the age of three years. An MRI ruled out insulinoma in all cases. One of these children had a history of a seizure prior to the diagnosis of CHI. A second patient reported episodes of weakness and pallor that resolved with sugar in early childhood. The third patient had no history suggestive of undiagnosed hypoglycemia.

### Genetics

A disease-causing variant was identified in 39% of those tested (n=19/49). All 19 children had persistent CHI. Variants were found in the *ABCC8* (n=13), *GLUD1* (n=2), *INSR* (n=1) and *HK1* (n=1) genes. In two patients a large deletion on the X chromosome or chr20p11.2 region was identified (Table 1).

**Table 1.** Genetic and clinical data for 24 probands with congenital hyperinsulinism and variants in a known hyperinsulinism gene. All 19 individuals with a disease-causing variant had persistent CHI.

| Patient ID | Gene | Variant | Inheritance | 18F-DOPA/PET-CT performed? | Diazoxide Response | Surgery performed |
| --- | --- | --- | --- | --- | --- | --- |
| **Disease-causing Variants** |  |  |  |  |  |  |
| 1 | *ABCC8* | p.(Arg598Ter)<br>c.1792C>T | Paternal | Focal | NR | Yes |
| 2 | *ABCC8* | p.(Arg168His)/p.(Glu9Ter)<br>c.503G>A/c.25G>T | Bi-allelic | Not performed | R | No |
| 3 | *ABCC8* | p.(Arg934Ter)/p.(Arg1215Gln)<br>c.2800C>T/c.3644G>A | Bi-allelic | Not performed | NR | No |
| 4 | *ABCC8* | p.(Asn1481fs)/p.(Leu81Pro)<br>c.4440del/c.242T>C* | Bi-allelic | Diffuse | NR | No |
| 5<sup>†</sup> | *ABCC8* | p.(Gly1379Asp)<br>c.4136G>A | *De novo* | Diffuse | R | No |
| 6 | *ABCC8* | p.(Gly505Arg)<br>c.1513G>C | *De novo* | Not performed | R | No |
| 7 | *ABCC8* | p.(Ser1483Asn)<br>c.4448G>A | *De novo* | Not performed | NR | No |
| 8 | *ABCC8* | p.(Leu1171fs)<br>c.3512del | Paternal | Diffuse | NR | Yes |
| 9<sup>†</sup> | *ABCC8* | p.(Gly1479Arg)<br>c.4435G>A | Unknown | Diffuse | R | No |
| 10 | *ABCC8* | p.(Leu508Pro)<br>c.1523T>C | Maternal | Not performed | R | No |
| 11 | *ABCC8* | p.(Tyr179Ter)<br>c.536_539del | Paternal | Focal | NR | Yes |
| 12 | *ABCC8* | p.(Asp1193fs)<br>c.3577del | Paternal | Focal in head of pancreas | NR | No |
| 13<sup>†</sup> | *ABCC8* | p.?<br>c.3653+1G>A | Unknown | Focal | NR | Yes |
| 14 | *GLUD1* | p.(Ser498Leu)<br>c.1493C>T | *De novo* | Not performed | R | No |
| 15 | *GLUD1* | p.(Ile497Met)<br>c.1491A>G | Unknown | Not performed | R | No |
| 16 | *INSR* | p.(Arg1191Gln)<br>c.3572G>A | Maternal | Not performed | R | No |
| 17 | *HK1* | Chr10:71,108,645T>C | Maternal | Not performed | R | No |
| 18** | Chr20p11.2del | Chr20:19,507,014-22,525,896del | Maternal | Not performed | R | No |
| 19 | Turner<br>syndrome | ChrX monosomy | *De novo* | Diffuse | NR | Yes |
| **Variants of uncertain clinical significance** |  |  |  |  |  |  |
| 20<sup>†</sup> | *ABCC8* | p.(Gly716Cys)<br>c.2146G>T | Unknown | Diffuse | R | No |
| 21 | *ABCC8* | p.(Val802Asp)<br>c.2405T>A | Paternal | Diffuse | NR | Yes |
| 22<sup>†</sup> | *ABCC8* | p.(Glu1208Lys)<br>c.3622G>A | Unknown | Not performed | R | No |
| 23 | *KCNJ11* | p.(Arg206Cys)<br>c.616C>T | Maternal | Not performed | R | No |
| 24 | *KCNJ11* | p.(Tyr268His)<br>c.802T>C | Paternal | Not performed | R | No |

\* This individual has a paternally inherited pathogenic *ABCC8* variant and a maternally inherited variant currently classified as of uncertain clinical significance (p.(Leu81Pro), c.242T>C). \*\* Patient and genetic results previously reported in <sup>36</sup>. † Genetic testing performed at a commercial laboratory. R = responsive to diazoxide, NR= not responsive to diazoxide.

In seven children, the variant had arisen *de novo* or co-segregated with CHI in the family, consistent with a dominant variant (*ABCC8* n=4, *GLUD1* n=1, *HK1* n=1, chrX deletion n=1). Compound heterozygous *ABCC8* variants were identified in three probands. In four children, a paternally inherited recessive *ABCC8* variant was found, and in two children, the variant was inherited from an unaffected parent (*INSR* and chr20del). For three children, the inheritance could not be established as parental samples were unavailable (*GLUD1* n=1, *ABCC8* n=2). Additionally, in 5 children (10%, n=5/49), a variant of uncertain significance (VUS) was identified (*ABCC8* n=3, *KCNJ11* n=2) (Table 1).

Risk factors for PSHI were identified in 10 of the 30 individuals without a genetic diagnosis. This included 6 individuals with transient CHI and 4 individuals with persistent CHI.

### Pancreatic imaging and surgical treatment

Imaging of the pancreas in 17 individuals with persistent CHI demonstrated diffuse tracer uptake in 8, focal uptake in 8 and atypical uptake in 1 (Table 1). Four of the eight children with diffuse uptake had undergone genetic testing prior to imaging. In three of these cases, the genetic results were consistent with diffuse disease, while one child had a paternally inherited recessive variant predicting a focal lesion. One additional child with diffuse disease underwent genetic testing following imaging and surgery which confirmed Turner syndrome. The child with atypical uptake had undergone genetic testing, but no disease-causing variant was found.

Three of the children with diffuse disease and the child with atypical tracer uptake underwent near-total pancreatectomy due to poor response to diazoxide. A second surgery was required in two patients because of persistent hypoglycemia. One child developed diabetes immediately post-surgery, while the other developed chyloperitoneum and exocrine insufficiency.

In the focal group, 4 children had undergone genetic testing prior to imaging which had identified a heterozygous *ABCC8* variant (3 paternal, 1 unknown) (Table 1). Lesionectomy was performed in 7 of the 8 children. In the remaining case, imaging revealed the lesion was in the head of pancreas, this patient was managed with octreotide and frequent feeding. Two of the focal cases developed hyperinsulinism post-surgery, which responded to octreotide. Diabetes or exocrine insufficiency was not reported in any of these cases.

### Clinical characteristics according to persistence of CHI and responsiveness to diazoxide

Patients with persistent diazoxide-unresponsive CHI had higher birthweights than those with persistent diazoxide-responsive or transient CHI (median Z-score 2.2 SDS vs 0.4 SDS vs −0.6 SDS respectively, ANOVA p<0.001). Insulin levels were also higher at presentation of disease in those who were unresponsive to diazoxide (median insulin level: 22.7 mU/l vs 12.2 mU/l (diazoxide-responsive) vs 11.0 mU/l (transient CHI) ANOVA p=0.018). Among patients with persistent CHI, those responsive to diazoxide were older at presentation than those with diazoxide-unresponsive (median 30 days vs 1 day, Kruskal Wallis p=0.001), or transient CHI (median 30 days vs 1 day, Kruskal Wallis p<0.001).

## Discussion

We describe 70 probands diagnosed with CHI in Argentina, making this the largest study to date reporting on the diagnosis and management of CHI in South America.^28^

Transient CHI was present in 33% (23/70) of individuals. This was characterized by an early presentation and a good response to diazoxide without side effects. Unlike other studies, there was no male predominance in the transient CHI cohort (52% male).^3,9,29^ At least one PSHI risk factor was detected in 82% of the transient CHI cases which aligns with previous reports.^9,29,30^ Four individuals with persistent CHI also had PSHI risk factors, including one infant with CHI remission in late infancy, who was born small for gestational age following a pregnancy affected by preeclampsia.

Genetic testing was performed on 7 individuals with transient CHI, all of whom had risk factors for PSHI, but no disease-causing variants were identified. This is consistent with other studies, which have shown that a genetic diagnosis is unlikely in individuals with transient CHI.^9^ Interestingly, the child whose CHI remitted in late infancy had a paternally inherited *KCNJ11* VUS, raising the possibility that this may be acting as a phenotypic modifier, potentially causing a prolonged form of PSHI.

Genetic analysis is limited in Argentina, so for 43 families, genetic testing was conducted in the UK through the Open Hyperinsulinism Genes Project, which is funded by the global charitable organization Congenital Hyperinsulinism International.^31^ A genetic diagnosis was obtained for 19 of the 49 (39%) patients who received testing. This is slightly lower than reported in other large series and may reflect a higher proportion of transient CHI cases in the cohort.8,10,32-35

All individuals with a genetic diagnosis had persistent CHI, with *ABCC8* variants accounting for 68% of diagnoses (n= 13/19). No homozygous variants were identified, consistent with the absence of consanguinity in the cohort. In two individuals finding a large contiguous gene deletion confirmed a syndromic form of CHI.^36,37^ Neither of these patients exhibited syndromic features at CHI diagnosis, although additional symptoms were diagnosed later in childhood which were consistent with the syndrome.

Sixteen children with transient CHI and 31 with persistent CHI were treated with diazoxide. This included an individual with a compound heterozygous *ABCC8* variant (p.(Arg168His)/p.(Glu9Ter)), an uncommon feature of biallelic variants in this gene.^38-40^ Four individuals experienced adverse effects to diazoxide, which were managed by reducing the dose and/or introducing adjunct therapy and one child treated with octreotide developed gallstones, a recognized side effect of this drug.^41,42^

Pancreatic imaging was performed on 17 individuals, 9 of these had received a genetic diagnosis prior to imaging which had correctly predicted the histology in 8 cases (Table 1). In the remaining case with diffuse uptake of the tracer, the presence of a paternal heterozygous variant had predicted focal disease. For this individual it is possible that there is a second, undetected variant, on the maternal allele or the patient had a giant focal lesion that appeared like diffuse disease on imaging.^43^

18F-DOPA PET-CT became available in Argentina in 2014, making it more accessible than genetic testing. As a result, some patients who had not undergone genetic testing were scanned in an attempt to avert prolonged medical treatment. With charity-funded genetic testing now available, screening of *ABCC8* and *KCNJ11* should be prioritized for future diazoxide-unresponsive cases, in line with current guidelines.^1,44^

Seven individuals with a focal lesion underwent lesionectomy. While all were diazoxide-unresponsive at surgery, one had initially responded to diazoxide (10 mg/kg/day) after a 10-hour fast, but later lost response. Surgery was curative in 71% (5/7) of cases, which is slightly lower than other reports.^17^ The child with a focal lesion in the pancreatic head and the two others who remained hypoglycemic post-surgery were treated with octreotide, with good response, similar to previous studies.^45,46^ In two cases, the octreotide dose was later reduced, indicating decreased disease severity.^47^

This retrospective study has some limitations. First, clinical data were collected from the case notes of patients managed at different tertiary centers across Argentina, this highlighted variability in the approach to managing the condition and incomplete data in some cases. Additionally, not all children with persistent CHI underwent genetic testing, and family member samples were sometimes unavailable to assess inheritance. Ensuring access to genetic testing for these families will be crucial to help inform on recurrence risk.

In conclusion, our study highlights the clinical and genetic heterogeneity of CHI in Argentina and underscores some of the many challenges clinicians face in managing this complex condition.

## Data Availability

All available clinical data is provided in the manuscript. The raw sequencing data generated during the current study are not publicly available to preserve patient confidentiality. Variant call format (.vcf) files are available through collaboration to experienced teams working on approved studies examining the mechanisms, cause, diagnosis and treatment of diabetes and other beta cell disorders. Requests for collaboration will be considered by a steering committee following an application to the Genetic Beta Cell Research Bank (IRAS: 316050, https://www.diabetesgenes.org/current-research/genetic-beta-cell-research-bank/). Contact by email should be directed to Sarah Flanagan.

## Acknowledgements

SEF has a Wellcome Trust Senior Research Fellowship (Grant Number 223187/Z/21/Z). We are grateful to Congenital Hyperinsulinism International (a501(c)3 organization) who funded the genetic testing conducted in Exeter for individuals with hyperinsulinism in Argentina through the Open Hyperinsulinism Genes Project.

## Author contributions

ATS designed the study. ATS, GP, GB, JC, GA, MA, MS, AF, VF, MVF and SM recruited patients and analysed the clinical data. JJB, JALH and SEF performed molecular genetic analysis and/or interpretation of the resulting data. ATS, JJB and SEF prepared the draft manuscript. All authors contributed to the discussion of the results and to manuscript presentation. All authors read and approved the final manuscript.

## References

1. De Leon DD, Arnoux JB, Banerjee I, et al. International Guidelines for the Diagnosis and Management of Hyperinsulinism. Horm Res Paediatr. 2024;97(3):279–298.

2. Bailey MJ, Rout A, Harding JE, Alsweiler JM, Cutfield WS, McKinlay CJD. Prolonged transitional neonatal hypoglycaemia: characterisation of a clinical syndrome. J Perinatol. 2021;41(5):1149–1157.

3. Hoe FM, Thornton PS, Wanner LA, Steinkrauss L, Simmons RA, Stanley CA. Clinical features and insulin regulation in infants with a syndrome of prolonged neonatal hyperinsulinism. J Pediatr. 2006;148(2):207–212.

4. Yau D, Laver TW, Dastamani A, et al. Using referral rates for genetic testing to determine the incidence of a rare disease: The minimal incidence of congenital hyperinsulinism in the UK is 1 in 28,389. PLoS One. 2020;15(2):e0228417.

5. Lapidus D, De Leon DD, Thornton PS, et al. The Birth Prevalence of Congenital Hyperinsulinism: A Narrative Review of the Epidemiology of a Rare Disease. Horm Res Paediatr. 2024:1–8.

6. Hewat TI, Johnson MB, Flanagan SE. Congenital Hyperinsulinism: Current Laboratory-Based Approaches to the Genetic Diagnosis of a Heterogeneous Disease. Front Endocrinol (Lausanne). 2022;13:873254.

7. Zenker M, Mohnike K, Palm K. Syndromic forms of congenital hyperinsulinism. Front Endocrinol (Lausanne). 2023;14:1013874.

8. Kapoor RR, Flanagan SE, Arya VB, Shield JP, Ellard S, Hussain K. Clinical and molecular characterisation of 300 patients with congenital hyperinsulinism. Eur J Endocrinol. 2013;168(4):557–564.

9. Mannisto JME, Maria M, Raivo J, et al. Clinical and Genetic Characterization of 153 Patients with Persistent or Transient Congenital Hyperinsulinism. J Clin Endocrinol Metab. 2020;105(4).

10. Snider KE, Becker S, Boyajian L, et al. Genotype and phenotype correlations in 417 children with congenital hyperinsulinism. J Clin Endocrinol Metab. 2013;98(2):E355–363.

11. Stanley CA, Lieu YK, Hsu BY, et al. Hyperinsulinism and hyperammonemia in infants with regulatory mutations of the glutamate dehydrogenase gene. N Engl J Med. 1998;338(19):1352–1357.

12. Glaser B, Kesavan P, Heyman M, et al. Familial hyperinsulinism caused by an activating glucokinase mutation. N Engl J Med. 1998;338(4):226–230.

13. Wakeling MN, Owens NDL, Hopkinson JR, et al. Non-coding variants disrupting a tissue-specific regulatory element in HK1 cause congenital hyperinsulinism. Nat Genet. 2022;54(11):1615–1620.

14. de Lonlay P, Fournet JC, Rahier J, et al. Somatic deletion of the imprinted 11p15 region in sporadic persistent hyperinsulinemic hypoglycemia of infancy is specific of focal adenomatous hyperplasia and endorses partial pancreatectomy. J Clin Invest. 1997;100(4):802–807.

15. Damaj L, le Lorch M, Verkarre V, et al. Chromosome 11p15 paternal isodisomy in focal forms of neonatal hyperinsulinism. J Clin Endocrinol Metab. 2008;93(12):4941–4947.

16. Verkarre V, Fournet JC, de Lonlay P, et al. Paternal mutation of the sulfonylurea receptor (SUR1) gene and maternal loss of 11p15 imprinted genes lead to persistent hyperinsulinism in focal adenomatous hyperplasia. J Clin Invest. 1998;102(7):1286–1291.

17. Adzick NS, De Leon DD, States LJ, et al. Surgical treatment of congenital hyperinsulinism: Results from 500 pancreatectomies in neonates and children. J Pediatr Surg. 2019;54(1):27–32.

18. Otonkoski T, Nanto-Salonen K, Seppanen M, et al. Noninvasive diagnosis of focal hyperinsulinism of infancy with [18F]-DOPA positron emission tomography. Diabetes. 2006;55(1):13–18.

19. Arya VB, Senniappan S, Demirbilek H, et al. Pancreatic endocrine and exocrine function in children following near-total pancreatectomy for diffuse congenital hyperinsulinism. PLoS One. 2014;9(5):e98054.

20. Beltrand J, Caquard M, Arnoux JB, et al. Glucose metabolism in 105 children and adolescents after pancreatectomy for congenital hyperinsulinism. Diabetes Care. 2012;35(2):198–203.

21. Houghton JA, Banerjee I, Shaikh G, et al. Unravelling the genetic causes of mosaic islet morphology in congenital hyperinsulinism. J Pathol Clin Res. 2020;6(1):12–16.

22. Boodhansingh KE, Yang Z, Li C, et al. Localized islet nuclear enlargement hyperinsulinism (LINE-HI) due to ABCC8 and GCK mosaic mutations. Eur J Endocrinol. 2022;187(2):301–313.

23. Hussain K, Flanagan SE, Smith VV, et al. An ABCC8 gene mutation and mosaic uniparental isodisomy resulting in atypical diffuse congenital hyperinsulinism. Diabetes. 2008;57(1):259–263.

24. Ellard S, Lango Allen H, De Franco E, et al. Improved genetic testing for monogenic diabetes using targeted next-generation sequencing. Diabetologia. 2013;56(9):1958–1963.

25. Laver TW, De Franco E, Johnson MB, et al. SavvyCNV: Genome-wide CNV calling from off-target reads. PLoS Comput Biol. 2022;18(3):e1009940.

26. Richards S, Aziz N, Bale S, et al. Standards and guidelines for the interpretation of sequence variants: a joint consensus recommendation of the American College of Medical Genetics and Genomics and the Association for Molecular Pathology. Genet Med. 2015;17(5):405–424.

27. Durkie M, Cassidy EJ, Berry I, Owens M, Turnbull C, Scott RH, Taylor RW, Deans ZC, Ellard S, Baple EL, McMullan DJ. ACGS Best Practice Guidelines for Variant Classification in Rare Disease 2024 2024.

28. Liberatore RDRJ, Monteiro ICM, Pileggi FO, Canesin WC, Sbragia L. Congenital hyperinsulinism and surgical outcome in a single tertiary center in Brazil. J Pediatr (Rio J). 2024;100(2):163–168.

29. Falzone N, Harrington J. Clinical Predictors of Transient versus Persistent Neonatal Hyperinsulinism. Horm Res Paediatr. 2020;93(5):297–303.

30. Hoermann H, Roeper M, Salimi Dafsari R, et al. Challenges in management of transient hyperinsulinism - a retrospective analysis of 36 severely affected children. J Pediatr Endocrinol Metab. 2021;34(7):867–875.

31. Raskin J, Pasquini TLS, Bose S, Tallis D, Schmitt J. Congenital Hyperinsulinism International: A Community Focused on Improving the Lives of People Living With Congenital Hyperinsulinism. Front Endocrinol (Lausanne). 2022;13:886552.

32. Demirbilek H, Arya VB, Ozbek MN, et al. Clinical characteristics and phenotype-genotype analysis in Turkish patients with congenital hyperinsulinism; predominance of recessive KATP channel mutations. Eur J Endocrinol. 2014;170(6):885–892.

33. Senniappan S, Sadeghizadeh A, Flanagan SE, et al. Genotype and phenotype correlations in Iranian patients with hyperinsulinaemic hypoglycaemia. BMC Res Notes. 2015;8:350.

34. Guven A, Cebeci AN, Ellard S, Flanagan SE. Clinical and Genetic Characteristics, Management and Long-Term Follow-Up of Turkish Patients with Congenital Hyperinsulinism. J Clin Res Pediatr Endocrinol. 2016;8(2):197–204.

35. Velde CD, Molnes J, Berland S, Njolstad PR, Molven A. Clinical and Genetic Characteristics of Congenital Hyperinsulinism in Norway: A Nationwide Cohort Study. J Clin Endocrinol Metab. 2025;110(2):554–563.

36. Laver TW, Wakeling MN, Caswell RC, et al. Chromosome 20p11.2 deletions cause congenital hyperinsulinism via the loss of FOXA2 or its regulatory elements. Eur J Hum Genet. 2024;32(7):813–818.

37. Gibson CE, Boodhansingh KE, Li C, et al. Congenital Hyperinsulinism in Infants with Turner Syndrome: Possible Association with Monosomy X and KDM6A Haploinsufficiency. Horm Res Paediatr. 2018;89(6):413–422.

38. Hewat TI, Yau D, Jerome JCS, et al. Birth weight and diazoxide unresponsiveness strongly predict the likelihood of congenital hyperinsulinism due to a mutation in ABCC8 or KCNJ11. Eur J Endocrinol. 2021;185(6):813–818.

39. Arya VB, Aziz Q, Nessa A, Tinker A, Hussain K. Congenital hyperinsulinism: clinical and molecular characterisation of compound heterozygous ABCC8 mutation responsive to Diazoxide therapy. Int J Pediatr Endocrinol. 2014;2014(1):24.

40. Martinez R, Fernandez-Ramos C, Vela A, et al. Clinical and genetic characterization of congenital hyperinsulinism in Spain. Eur J Endocrinol. 2016;174(6):717–726.

41. Demirbilek H, Shah P, Arya VB, et al. Long-term follow-up of children with congenital hyperinsulinism on octreotide therapy. J Clin Endocrinol Metab. 2014;99(10):3660–3667.

42. Herrera A, Vajravelu ME, Givler S, et al. Prevalence of Adverse Events in Children With Congenital Hyperinsulinism Treated With Diazoxide. J Clin Endocrinol Metab. 2018;103(12):4365–4372.

43. Ismail D, Kapoor RR, Smith VV, et al. The heterogeneity of focal forms of congenital hyperinsulinism. J Clin Endocrinol Metab. 2012;97(1):E94–99.

44. Banerjee I, Skae M, Flanagan SE, et al. The contribution of rapid KATP channel gene mutation analysis to the clinical management of children with congenital hyperinsulinism. Eur J Endocrinol. 2011;164(5):733–740.

45. Mannisto JME, Jaaskelainen J, Otonkoski T, Huopio H. Long-Term Outcome and Treatment in Persistent and Transient Congenital Hyperinsulinism: A Finnish Population-Based Study. J Clin Endocrinol Metab. 2021;106(4):e1542–e1551.

46. Arya VB, Guemes M, Nessa A, et al. Clinical and histological heterogeneity of congenital hyperinsulinism due to paternally inherited heterozygous ABCC8/KCNJ11 mutations. Eur J Endocrinol. 2014;171(6):685–695.

47. Salomon-Estebanez M, Flanagan SE, Ellard S, et al. Conservatively treated Congenital Hyperinsulinism (CHI) due to K-ATP channel gene mutations: reducing severity over time. Orphanet J Rare Dis. 2016;11(1):163.

